# Time-aggregated mobile phone mobility data are sufficient for modelling influenza spread: the case of Bangladesh

**DOI:** 10.1101/2020.03.11.20033555

**Authors:** Solveig Engebretsen, Kenth Engø-Monsen, Mohammad Abdul Aleem, Emily Suzanne Gurley, Arnoldo Frigessi, Birgitte Freiesleben de Blasio

## Abstract

Human mobility plays a major role in the spatial dissemination of infectious diseases. We develop a spatio-temporal stochastic model for influenza-like disease spread based on estimates of human mobility. The model is informed by mobile phone mobility data collected in Bangladesh. We compare predictions of models informed by daily mobility data (reference) with that of models informed by time-averaged mobility data, and mobility model approximations. We find that the gravity model overestimates the spatial synchrony, while the radiation model underestimates the spatial synchrony. Using time-averaged mobility resulted in spatial spreading patterns comparable to the daily mobility model. We fit the model to 2014-2017 influenza data from sentinel hospitals in Bangladesh, using a sequential version of Approximate Bayesian Computation. We find a good agreement between our estimated model and the case data. We estimate transmissibility and regional spread of influenza in Bangladesh, which are useful for policy planning. Time-averaged mobility appears to be a good proxy for human mobility when modelling infectious diseases. This motivates a more general use of the time-averaged mobility, with important implications for future studies and outbreak control. Moreover, time-averaged mobility is subject to less privacy concerns than daily mobility, containing less temporal information on individual movements.

## 1 Introduction

Mathematical models are an essential tool to understand and predict epidemic spread in space and time (1). Human mobility is a main driver for the spatial dissemination of infectious diseases. It is therefore pivotal to include a sensible model for human movement in spatial disease models.

The gravity model (2) is the most widely used model for human mobility. It assumes that the flux of movements between locations increases with population sizes and decays with distance. The recently proposed radiation model (3) puts more emphasis on the population density between the locations, not only their distance. The models have primarily been developed and assessed for developed countries (4), with use of commuting and long-range travel. However, little data have been published on the transportation and commuting networks in developing countries.

In recent years, there has been a surge in the availability of large data on human movements. For example, exploiting telecommunication data allows mobility observations of large populations with high resolution in time and space. Mobile phone data have previously been used to improve understanding of infectious disease spread, see e.g. (4; 5; 6). In contrast, census data on human mobility offer a snapshot of the mobility behaviour, and are subject to recall bias and limitation in size (7).

Because mobile phone mobility data contain rich location data about individuals, they are subject to privacy challenges (8). In practice, it is often difficult to get access to mobile phone data for research purposes, privacy concern being a major hindrance. Use of time-aggregated mobile phone data provides a means to protect peoples’ identities. Additionally, such data are not specific to calendar dates, and therefore they may have a broader use outside the time period where they were collected. However, until now there have been few attempts to compare, in a coherent way, the use of mobile phone data with different time resolution to predict infectious disease dynamics. Not least for developing countries, where human mobility data are scarce. Understanding the limitations of using time-averaged mobile phone data and model approximations to human movement is essential to guide the choice of mobility measures in models and further development in this field. Additionally, mathematical models are important for use in public health emergency planning.

Bangladesh is a suitable study setting to address these questions. The country does not have detailed census data for commuting and travel flow prediction, and synthetic models for movement patterns or mobile phone data are therefore in demand. Bangladesh belongs to the group of least developed countries in the world, and respiratory infections are among the leading causes of death. Seasonal influenza has been estimated to cause an estimated 6097 and 16804 deaths in 2010 and 2011 (9), with a total cost of about US$ 169 million in 2010 (10). Moreover, the country is a likely source of novel avian influenza viruses capable of causing pandemics (11). Timely modelling of influenza outbreaks is important for public health pandemic preparedness planning.

Here, we conduct a data-driven simulation study to compare the spatial dissemination of influenza in Bangladesh using highly detailed mobile phone data. To this aim, we extend a fine-scaled stochastic *SEII*_*a*_ *R* metapopulation model developed in (12), and fit the model to influenza hospital case data. We integrate the model with different mobility approximations, and investigate the spatial transmission for each model at different geographic resolutions, and for different location-specific seeds.

We demonstrate that time-averaged mobile phone mobility data capture well the disease spreading pattern of daily mobile phone data. Exploring synthetic mobility models, we show that the gravity model produces consistent outcomes at a global scale, but has poor ability to predict disease spread at lower spatial granularity. The radiation model predicted overall delayed and too heterogeneous disease spreading pattern. Using hospital sentinel data from 2014-2017, we provide novel evidence for the transmissibility of seasonal influenza in a developing country. Finally, we document the feasibility of applying sequential Monte Carlo Approximate Bayesian Computation (ABC-SMC)-techniques to estimate parameters in a stochastic metapopulation model informed by scarce influenza case data.

## 2 Data

### 2.1 Mobile phone data

The mobile phone data are provided by Telenor, through its subsidiary Grameenphone. The data are aggregated and anonymised, and contain movement information for 60 million customers throughout the country (*≈* 37% of the population) from 1. April 2017 to 30. September 2017 (183 calendar days), covering the typical influenza season. Bangladesh is divided into seven administrative units (Figure 1), which are further divided into 64 districts and 544 subdistricts called upazilas.

**Figure 1:**
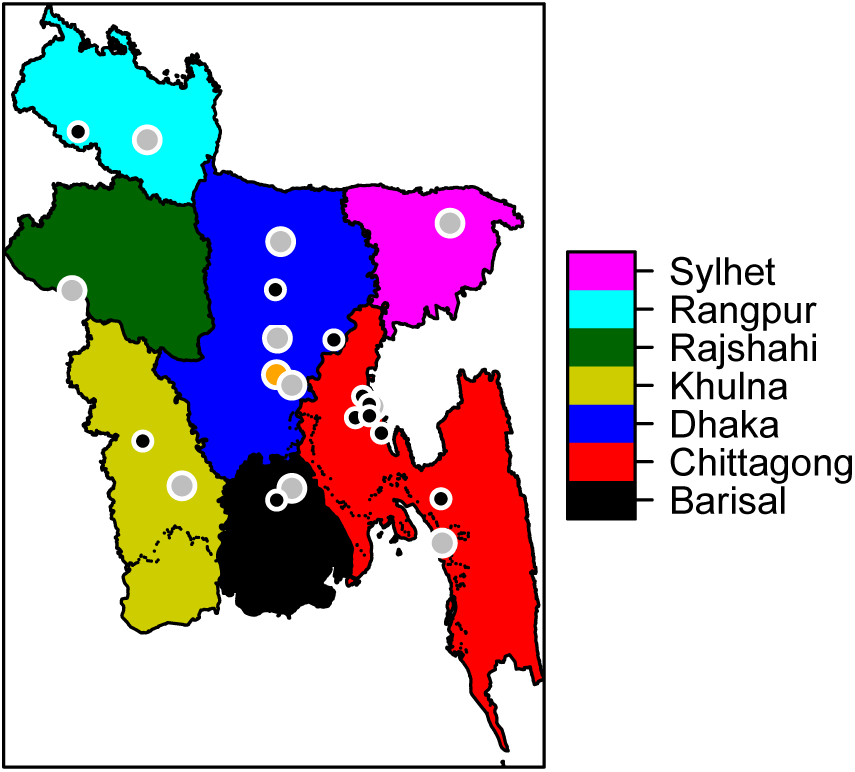
The divisions of Bangladesh. The city corporations are marked by grey points, while Dhaka city, the capital, is marked by an orange point. The black points are the earliest detected influenza cases for 2017.

Whenever a phone call is made or a text message is sent, it is routed through the nearest cell tower. Each day, users are assigned to their most frequent cell tower location. This provides a time series of locations for each user. The mobile phone data are aggregated into 183 daily mobility matrices at upazila level, by counting the subscribers who have transitioned between upazilas, or remained in the same location, from one day to the next. We do not have individual identifiers in the mobility data. We use the transition matrices to estimate the population size in each upazila, by scaling the average number of subscribers to the total population of Bangladesh: 163 million as of 2016 (13). The mobility matrices are scaled up to match the estimated population sizes in each upazila. In addition, we calculate one time-averaged mobility matrix for the entire study period. A few upazilas are missing in the mobility data, and left out from the model (see electronic supplementary material section S1 for details).

The estimated mean daily proportion travelling between upazilas is 0.278, with standard deviation (std) 0.0148, indicating little variation between days in the proportion travelling. The estimated daily proportion of the population travelling between upazilas displays weekly cycles and marked dips and peaks during major holidays (Figure 2).

**Figure 2:**
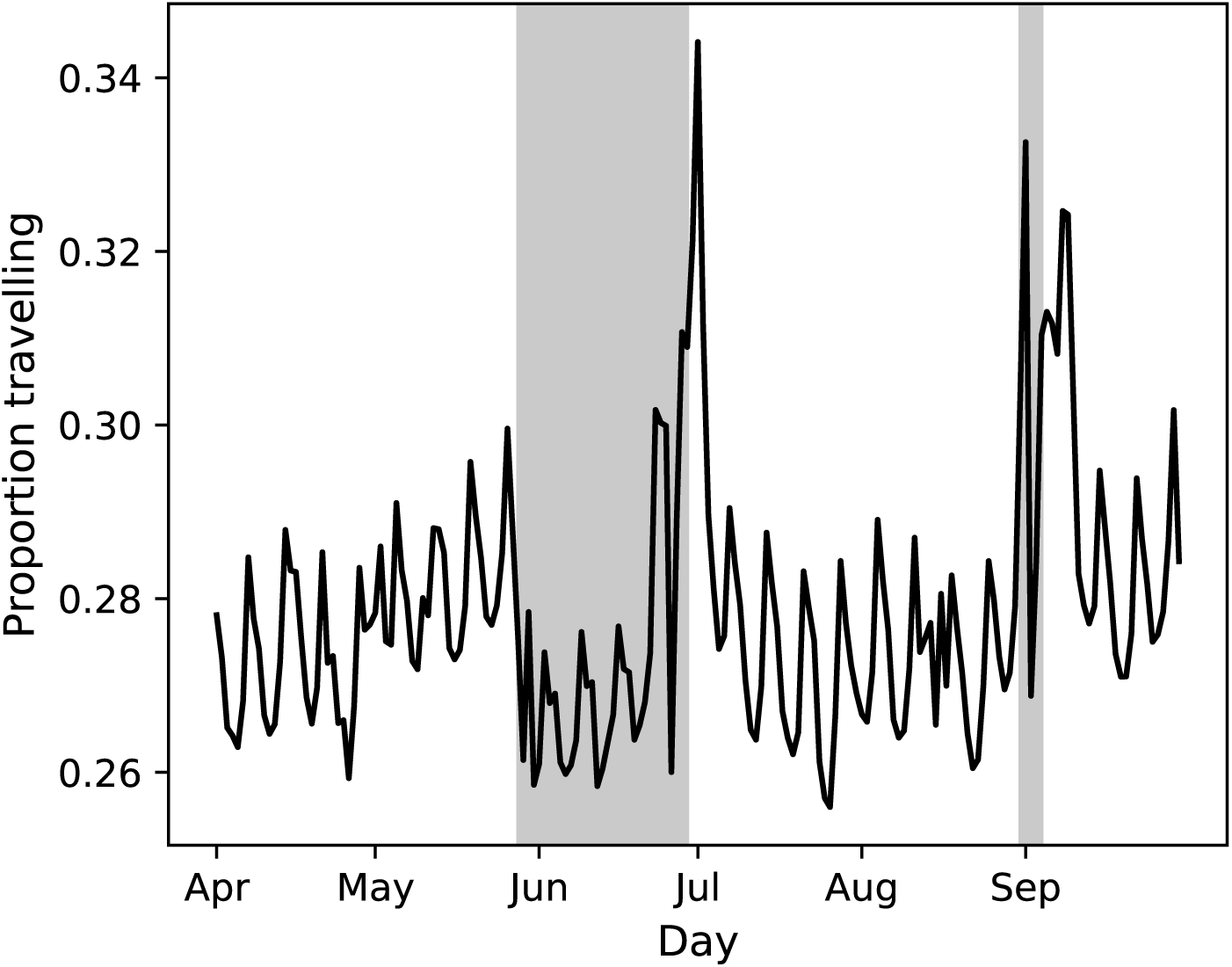
Estimated proportion of population travelling between upazilas each day. Eid al-Fitr occurred on June 25th-26th, Eid al-Adha occurred on September 1st-2nd, and Ramadan on May 26th-June 24th.

The most popular travelling routes are connected to major cities, and around each there is a star-like flow (Figure 3). The mobility network is dense. The density of the time-averaged mobility (the ratio of existing links to potential links) is 82%. The average density of the daily networks is 42% (std 3.1%). More network statistics are provided in the electronic supplementary material Section S16.

**Figure 3:**
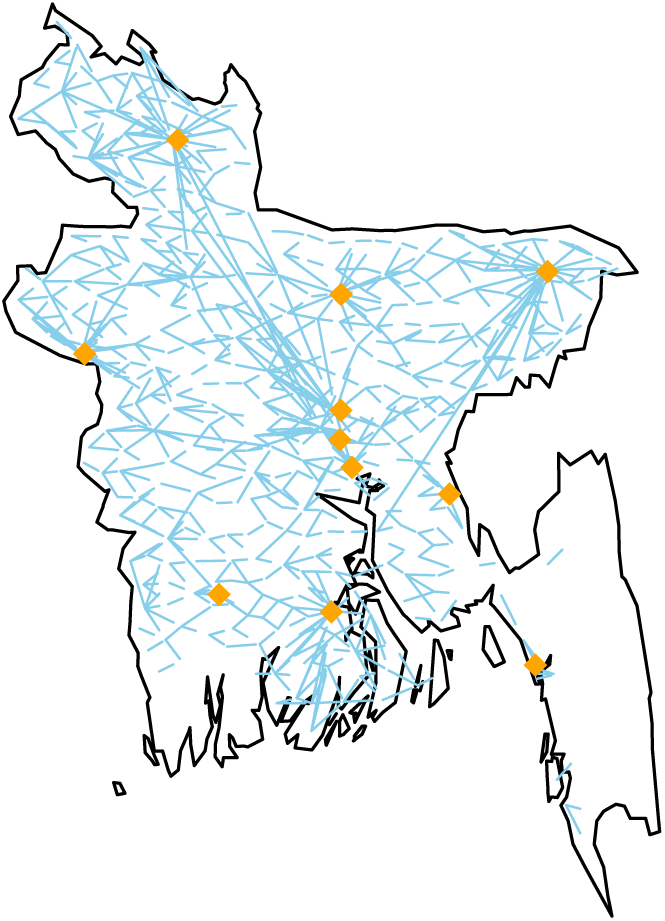
The mobility routes with more than 2000 travellers daily. The city corporations are marked.

### 2.2 Influenza data

The influenza data are provided by International Centre for Diarrhoeal Disease Research, Bangladesh. The data contain daily laboratory-confirmed influenza cases from 12 sentinel hospitals, covering all seven divisions of Bangladesh.

The patients eligible for testing are the inpatients with severe acute respiratory infection (SARI, inpatients aged 5 years and above), severe pneumonia (inpatients younger than 5 years), and outpatients with influenza-like illness. From July 2017, the SARI case definition was used for all ages. The total number of influenza cases for 2017 is 890, out of 4229 tested. We use patient residence information to obtain observed case counts in each upazila. The weekly incidence for the 2017 seasonal influenza shows a dip right before the unimodal peak at the June-July transition. This is likely an artefact caused by limited testing activity and/or healthcare seeking during the holidays. More descriptive statistics for the case data for all seasons considered, 2014-2017, are provided in electronic supplementary material Section S2.

## 3 Models and methods

The influenza spread model is a closed metapopulation model on upazila level (14; 15) for Bangladesh, with a local stochastic disease process in every upazila. The local transmission processes are coupled through individuals who travel, according to mobility estimates.

### 3.1 Infectious disease model

To model the influenza dynamics within each upazila, a stochastic compartmental *SEII*_*a*_ *R* model is used. The detailed model description, equations, parameter definitions, and values are provided in electronic supplementary material Section S3.

Let *w*_*i,j,t*_ denote the number of travellers from location *i* to location *j* on day *t*, estimated from the mobile phone data. For days that are not covered by the mobile phone data (before April 1st 2017 and after September 30th 2017), we use the time-averaged mobility. The number of people in location *i* on calendar day *t*, 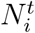, is given by

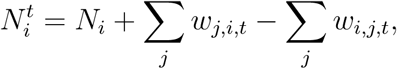

where *N*_*i*_ is the population size in location *i*. The travelling individuals are selected uniformly at random from the home population. The model is as follows: at the beginning of each day, the individuals travel according to the mobility estimates. Then they mix at their destination location for one day, before we send them back to their home location.

We assume that the number of observed cases on a day can be modelled as a binomial process, with the number of trials equal to the actual number of new symptomatic cases, and success probability *r*. Thus, *r* is the probability for a symptomatic influenza case to be reported in the hospital sentinel data. We estimate two parameters; the transmission rate parameter *β*, which is directly related to the effective reproductive number, *R*_*e*_, and the reporting probability *r*, which is related to the severity of the influenza season. In severe seasons, one expects more hospitalised cases, and thus a higher reporting probability than for milder influenza seasons.

We estimate the parameters using approximate likelihoods based on model simulations, through ABC-SMC (16), using the 2017 case data. The idea is based on a Monte Carlo scheme, where the accepted parameters are such that the simulated epidemic with these parameters is sufficiently close to the observed data, measured in summary statistics. We choose four summary statistics: duration, final size, peak height, and global incidence curve. The ABC-SMC algorithm and details on the summary statistics are provided in the electronic supplementary material Section S6. The algorithm is similar to that used in (17).

### 3.2 Mobility models

Instead of using the mobile phone movement data, one could assume certain classical models. We compare the usefulness of the telecommunication data with a gravity model (2) and the radiation model (3).

The estimated number of travellers between locations *i* and *j* for the gravity model, 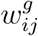, and the radiation model, 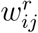, are given by

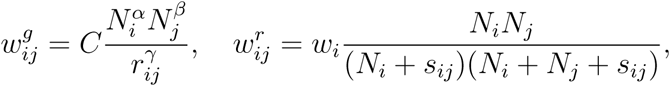

where *N*_*i*_ and *N*_*j*_ are the population sizes in locations *i* and *j*, respectively, *r*_*ij*_ is the distance between the locations, *α, β, γ* and *C* are adjustable parameters, found by fitting the gravity model to the time-averaged mobility, *s*_*ij*_ denotes the total population in the circle of radius *r*_*ij*_, centred at location *i*, and *w*_*i*_ is the total number of travellers from location *i*. We assume that *w*_*i*_ is proportional to population size as in (3), and use the time-averaged mobility to estimate the total number of travellers.

The estimated gravity model is

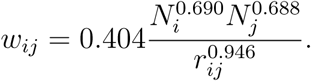

### 3.3 Simulation set-up, seeding, and model outcomes

To simulate the 2017 influenza season using the *SEII*_*a*_ *R* model, we use the posterior modes from ABC-SMC as point estimates for *β* and *r*. These parameter estimates are used for all simulations, except the 2014-2016 simulations, where the parameters are tuned to match the case data for these seasons.

We choose as outcome measures: initial date, final size, peak date, and peak prevalence. The initial date is defined as the date where, for seven consecutive days, the prevalence in the location exceeds 0.1% of the population. We compare initial dates on upazila and division level. The final size is defined as the total number of symptomatic infections, the peak date is the date with the largest number infected, and the peak prevalence is the proportion infected on the peak date. These are compared on national level. As the process is stochastic, we perform 100 simulations in each setting and compare the averages.

We seed the epidemic with ten infectious individuals in each of the eleven upazilas with earliest confirmed influenza cases in 2017 (Figure 1). The seeding date (set to day 0) is tuned to match the observed peak in the case data. When investigating spatial spread with different seeding locations, the same total number of infectious cases (110) is placed in a single upazila of high population density in the respective division. To simulate the 2014-2016 influenza seasons, we seed in the eleven upazilas with the earliest cases for these seasons, and tune the seeding date to match the case data. Details on the seeding locations are provided in electronic supplementary material Section S7.

Four different mobility proxies are considered: 1) daily mobility data 1st April 2017-30th September 2017, 2) 6-month averaged daily mobility data, “time-averaged mobility”, 3) gravity model informed by time-averaged mobility, and 4) radiation model. In addition, in order to assess the advantage of having daily mobility in the initial period, we also seed on the 1st of April 2017 using the daily mobility data, with the same seeding scenarios.

## 4 Results

### 4.1 Model fit for 2017 seasonal influenza

We estimated an *R*_*e*_ of 1.220 (Table 1, first row). The best match between the peak in the influenza data and the model was obtained by seeding on the 30th September 2016. The posterior distributions of *β* and *r* were both unimodal (Figure 5). The estimated *r* suggests that approximately 4 per 100 000 infectious cases in Bangladesh were detected in the hospital sentinel data. The overall shape of the simulated incidence curve fits well the up-scaled hospital data in accordance with the estimated reporting probability (Figure 6), but the case data are spiky due to the limited case numbers and variation in testing activity. The correlation between the simulations and the case data was 0.63.

**Table 1:** Parameter estimates.

| Year | $R_e$ | $\beta$ | $r$ |
| --- | --- | --- | --- |
| 2017 | 1.220 (1.201, 1.237) | 0.487 (0.479, 0.494) | $3.52 \cdot 10^{-5}$ ($5.02 \cdot 10^{-6}$, $8.34 \cdot 10^{-5}$) |
| 2016 | 1.196 (1.180, 1.211) | 0.478 (0.471, 0.484) | $2.15 \cdot 10^{-5}$ ($2.75 \cdot 10^{-6}$, $8.07 \cdot 10^{-5}$) |
| 2015 | 1.152 (1.141, 1.164) | 0.460 (0.455, 0.465) | $2.49 \cdot 10^{-5}$ ($3.58 \cdot 10^{-6}$, $1.10 \cdot 10^{-4}$) |
| 2014 | 1.213 (1.198, 1.235) | 0.484 (0.478, 0.493) | $2.89 \cdot 10^{-5}$ ($3.65 \cdot 10^{-6}$, $7.74 \cdot 10^{-5}$) |

Posterior modes for $R_e$ , $\beta$ and $r$ for different seasons. 95% posterior credibility intervals are given in parenthesis.

**Figure 4:**
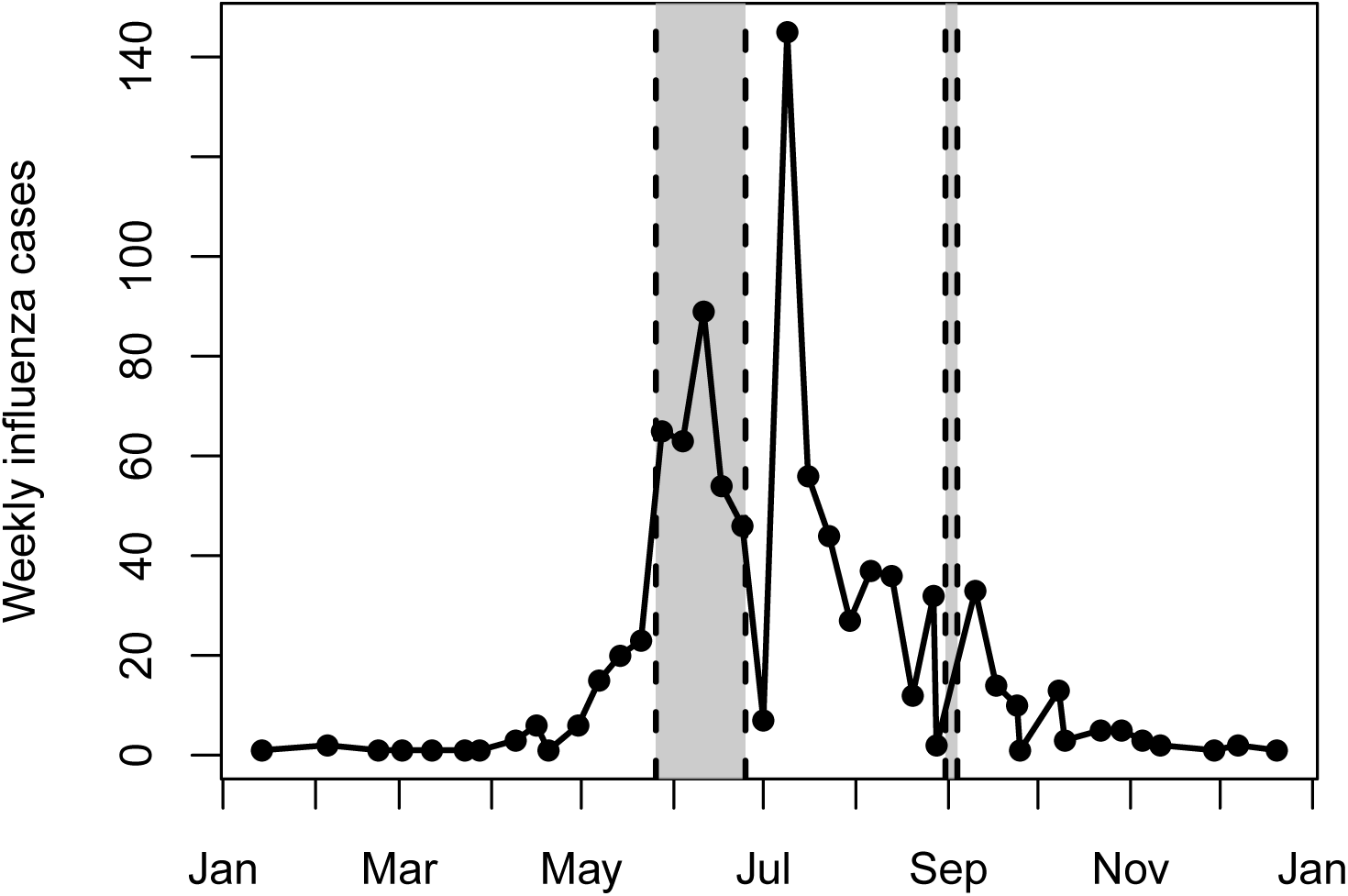
Weekly incidence of positive influenza cases in hospital sentinel data from Bangladesh, 2017.

**Figure 5:**
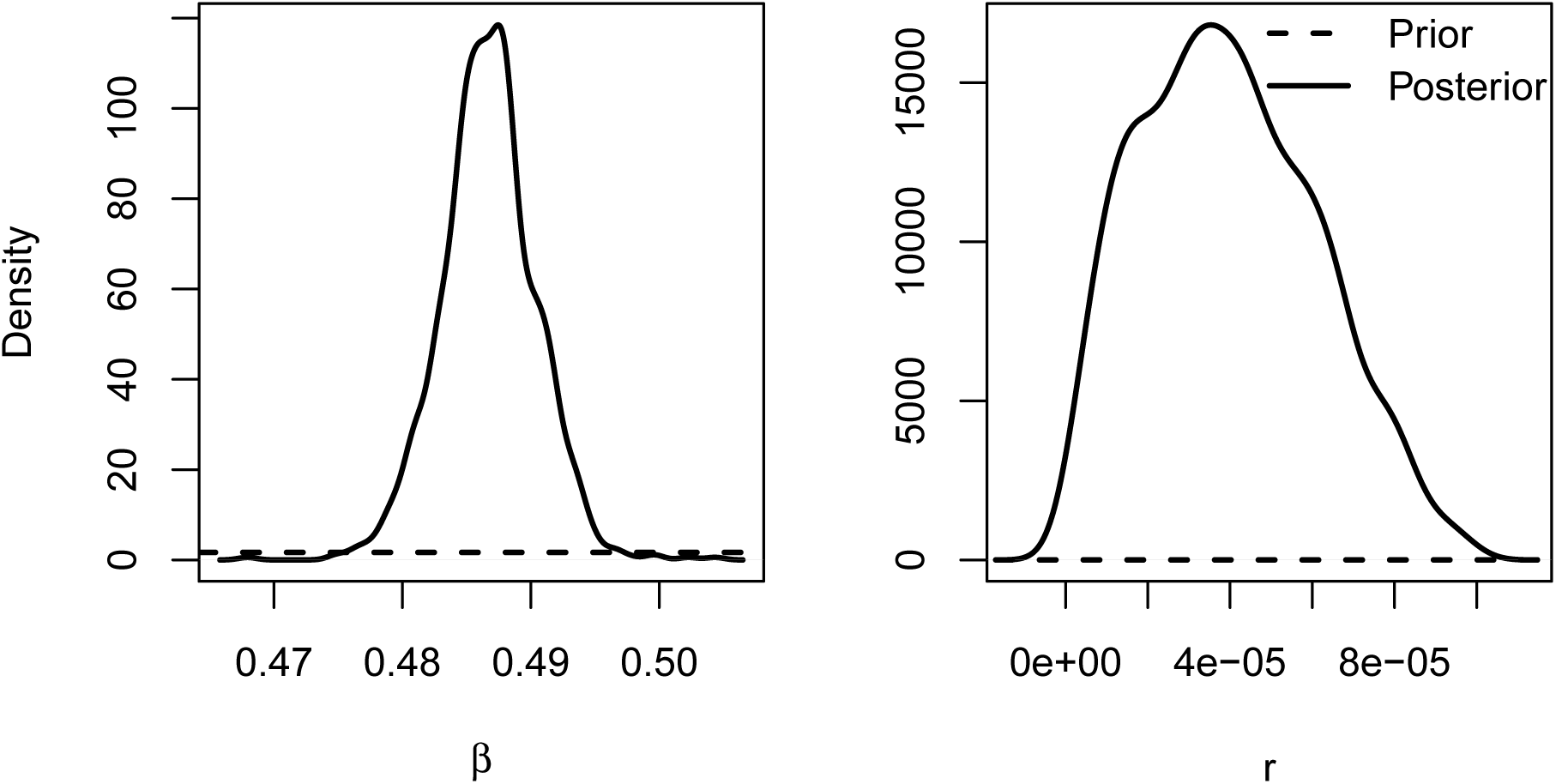
Posterior distribution of *β* and *r* for 2017.

**Figure 6:**
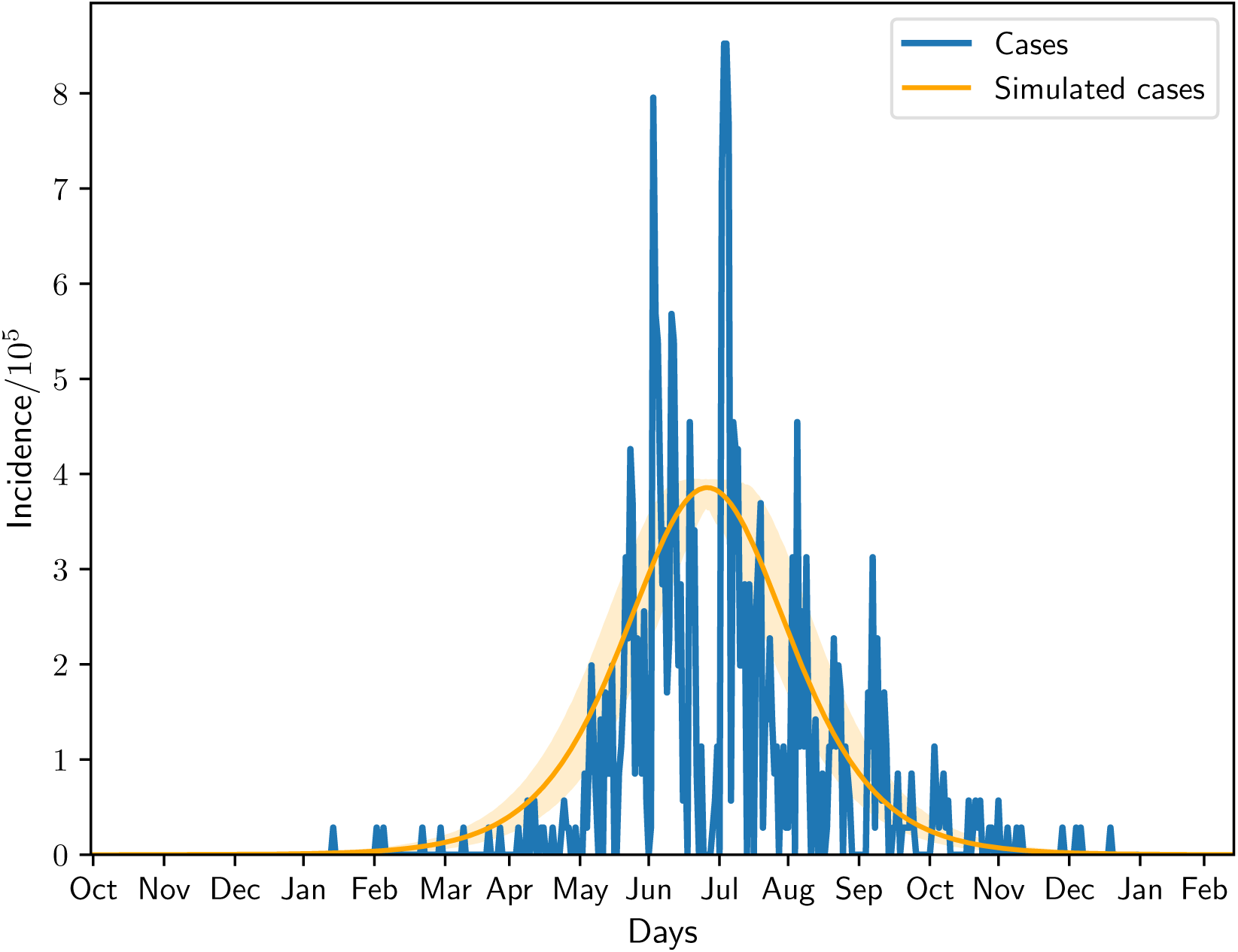
Simulated mean incidence for the 2017 seasonal influenza with 95% confidence interval, and scaled case data according to the estimated *r*.

### 4.2 Comparison of mobility models

We evaluated the disease-spreading pattern of the metapopulation model integrated with different mobility approximations, by their deviation from outcomes of the model informed by daily mobility, henceforward named “daily mobility model”. We compared the model performances at different geographic resolution, and for different seeding scenarios, including the early hospital cases in 2017, and division-specific seed.

At national scale, the daily mobility model predicted a final size of about 0.23 of symptomatic infections in the Bangladeshi population, independent of seeding (Table 2, top). The peak date of the 2017 epidemic was about 272 days, similar to that obtained by a seed in Dhaka or Chittagong (cf. Figure 1). Seeding in the other divisions delayed the peak by 1-5 days. The peak prevalence was roughly 0.7%, and was relatively constant across all seeding scenarios. The time-averaged mobility model gave similar output (Table 2, middle-top). The gravity model produced almost constant outputs for all seeds, with 2-7 days earlier peak dates and a slightly larger peak compared to the daily mobility model (Table 2, middle-bottom). The radiation model predicted similar and constant final sizes (Table 2, bottom), delayed peak dates by 1-5 weeks, while it underestimated the peak prevalence on the order of 5-20% compared to the reference.

**Table 2:**
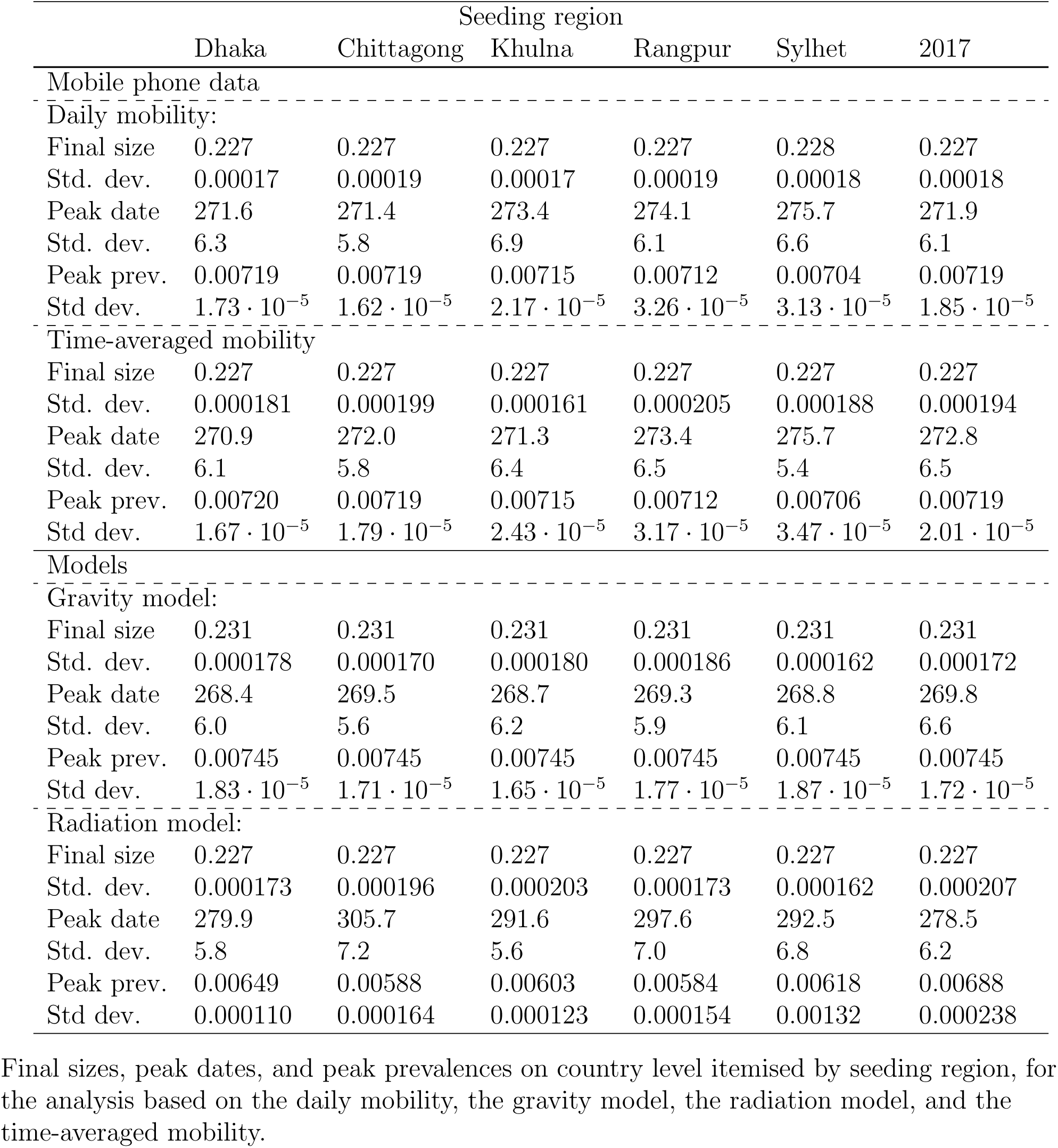
Global final size, peak date, and peak prevalence (N = 100 simulations).

At a finer spatial scale, we compared the mean initial date distribution at the division level, and by seed region. For the daily mobility model (Figure 7), the distributions were very overlapping when seeding in the most densely populated regions, while the spread was less coherent for other seeding scenarios.

**Figure 7:**
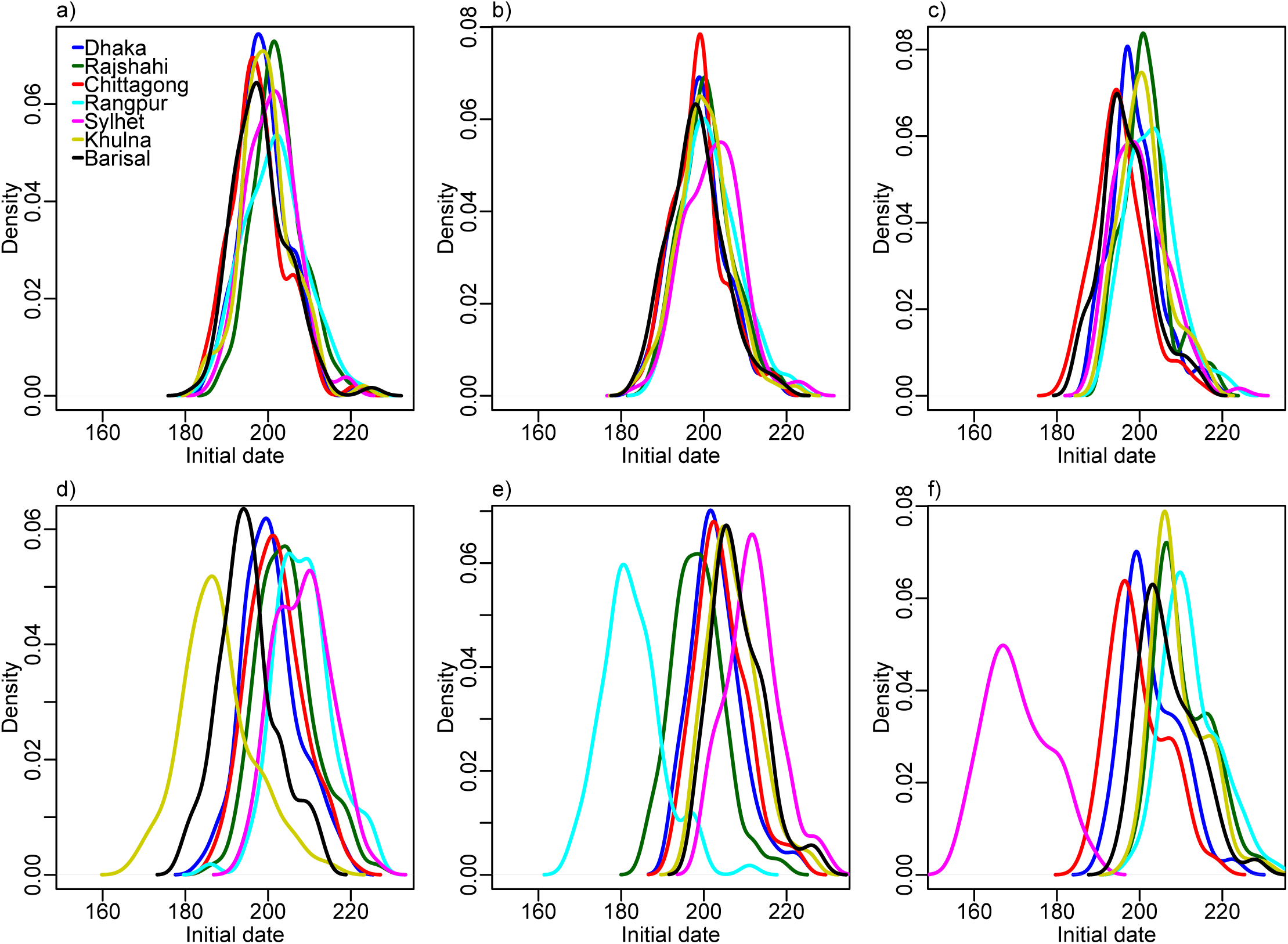
Daily mobility model. Initial dates distributions when seeding in a) 2017 simulation, b) Dhaka, c) Chittagong, d) Khulna, e) Rangpur, and f) Sylhet.

The initial dates distributions for the gravity model, the radiation model, the time-averaged mobility, and the daily mobility when seeding on the 1st of April 2017 are provided in Figures 8-11, respectively. When seeding on the 30th of September 2016, we had daily mobility for the period with most epidemic activity. When seeding on the 1st of April 2017, we had daily mobility in the start of the epidemic. The initial dates distributions indicate overestimation of the spatial synchrony for the gravity model, and underestimation of the spatial synchrony for the radiation model, by comparing the overlap of the distributions for the different divisions and seeding scenarios. For the time-averaged mobility, the initial dates distributions were similar to both seeding dates with daily mobility.

**Figure 8:**
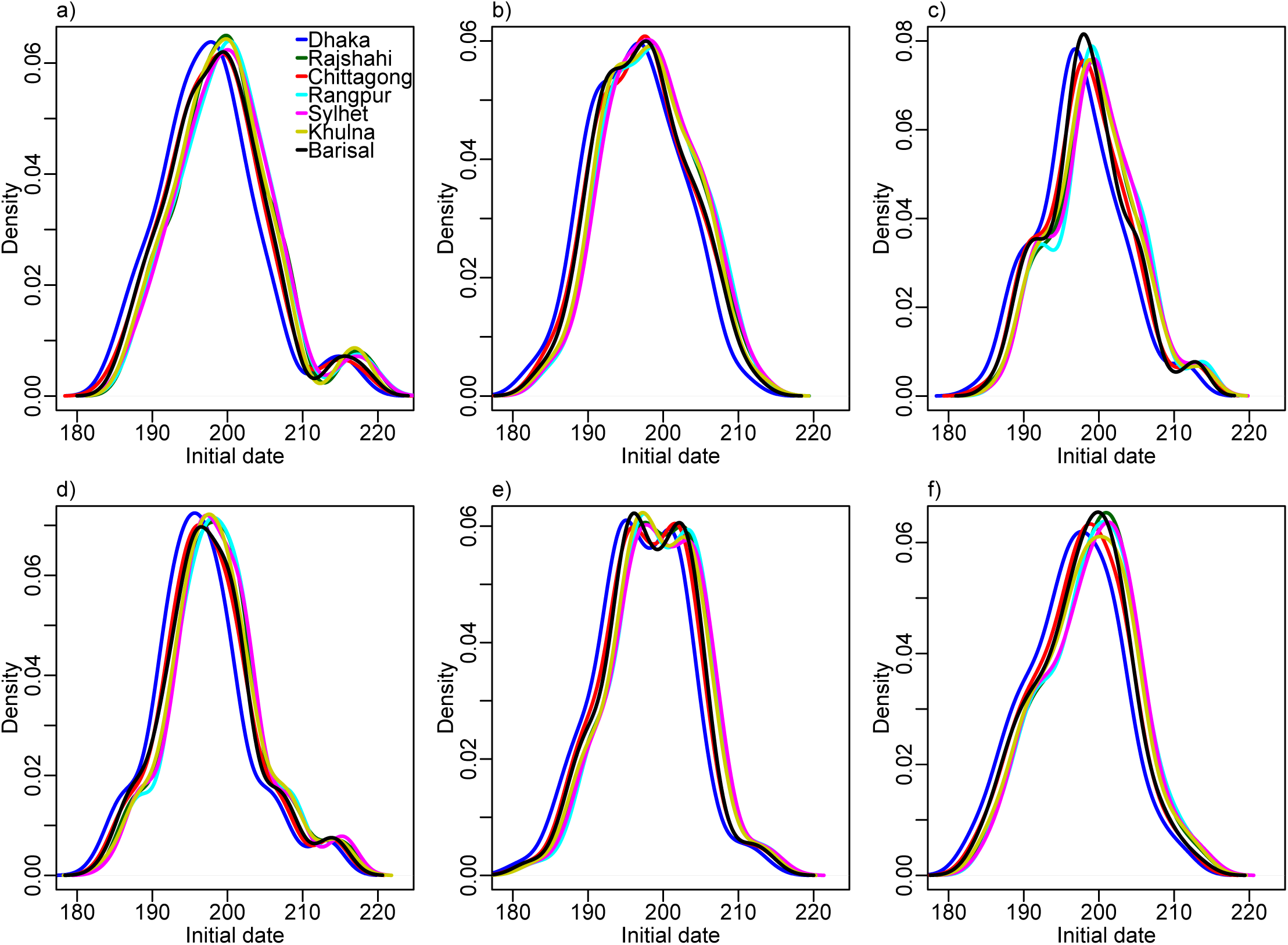
Gravity model. Initial dates distribution for each division for the gravity model, seeding in a) 2017 simulation, b) Dhaka, c) Chittagong, d) Khulna, e) Rangpur, and f) Sylhet.

The orders of the mean initial date for each division are provided in Table 3. The results with the gravity model give a false impression of the spatial spread being robust to seeding location. For the radiation model, the earliest initial date was consistently for the seeding division. For the time-averaged mobility, the ordering was similar to that of the daily mobility, for both seeding dates. In particular, the relative arrival times were identical for four of the seeding scenarios, with minor variations for the rest.

**Table 3:** Order of arrival (N = 100 simulations).

| Seeding division | Dhaka | Chittagong | Khulna | Rangpur | Sylhet | Barisal | Rajshahi |
| --- | --- | --- | --- | --- | --- | --- | --- |
| Daily mobility: |  |  |  |  |  |  |  |
| Dhaka | 3 | 2 | 4 | 6 | 7 | 1 | 5 |
| Chittagong | 3 | 1 | 5 | 7 | 4 | 2 | 6 |
| Khulna | 3 | 4 | 1 | 7 | 6 | 2 | 5 |
| Rangpur | 3 | 4 | 5 | 1 | 7 | 6 | 2 |
| Sylhet | 3 | 2 | 5 | 7 | 1 | 4 | 6 |
| 2017 | 3 | 1 | 4 | 6 | 5 | 2 | 7 |
| Gravity model: |  |  |  |  |  |  |  |
| Dhaka | 1 | 2 | 4 | 6 | 7 | 3 | 5 |
| Chittagong | 1 | 2 | 4 | 7 | 6 | 3 | 5 |
| Khulna | 1 | 2 | 4 | 7 | 6 | 3 | 5 |
| Rangpur | 1 | 2 | 4 | 7 | 6 | 3 | 5 |
| Sylhet | 1 | 2 | 4 | 7 | 7 | 6 | 5 |
| 2017 | 1 | 2 | 4 | 7 | 6 | 3 | 5 |
| Radiation model: |  |  |  |  |  |  |  |
| Dhaka | 1 | 2 | 5 | 7 | 3 | 4 | 6 |
| Chittagong | 4 | 1 | 3 | 7 | 5 | 2 | 6 |
| Khulna | 3 | 4 | 1 | 7 | 6 | 2 | 5 |
| Rangpur | 3 | 6 | 4 | 1 | 5 | 7 | 2 |
| Sylhet | 2 | 3 | 7 | 5 | 1 | 6 | 4 |
| 2017 | 3 | 1 | 5 | 6 | 4 | 2 | 7 |
| Time-averaged mobility: |  |  |  |  |  |  |  |
| Dhaka | 3 | 2 | 4 | 7 | 6 | 1 | 5 |
| Chittagong | 3 | 1 | 5 | 7 | 4 | 2 | 6 |
| Khulna | 3 | 4 | 1 | 7 | 6 | 2 | 5 |
| Rangpur | 3 | 4 | 5 | 1 | 7 | 6 | 2 |
| Sylhet | 3 | 2 | 5 | 7 | 1 | 4 | 6 |
| 2017 | 3 | 1 | 4 | 5 | 7 | 2 | 6 |
| Seeding 1st April 2017: |  |  |  |  |  |  |  |
| Dhaka | 3 | 1 | 4 | 6 | 7 | 2 | 5 |
| Chittagong | 3 | 1 | 5 | 7 | 4 | 2 | 6 |
| Khulna | 3 | 4 | 1 | 7 | 6 | 2 | 5 |
| Rangpur | 3 | 4 | 5 | 1 | 7 | 6 | 2 |
| Sylhet | 3 | 2 | 5 | 7 | 1 | 4 | 6 |
| 2017 | 3 | 1 | 4 | 5 | 6 | 2 | 7 |

Relative order of arrival in terms of mean initial dates for the different seeding scenarios, for the analysis based on daily mobility, gravity model, radiation model, time-averaged mobility, and on daily mobility when seeding on the 1st of April 2017. Each row gives the arrival order when seeding in different locations.

Finally, we compared the mobility approximations on the upazila level. The differences in initial dates between the mobility approximations and the daily mobility for each upazila are given in Figure 12, for the 2017 simulation setting. The gravity model resulted in slightly more negative values than positive, indicating too early initial dates (Figure 12a). The upazilas of the North-West experienced too early initial dates under the gravity model. The correlation between the mean initial dates on upazila level under the gravity model and the daily mobility model was 0.74. The upazilas in the South-East experienced too early initial dates under the radiation model, and the upazilas in the North-West experienced delayed initial dates (Figure 12b). The correlation between the mean initial dates under the radiation model and the daily mobility model was 0.69.

**Figure 9:**
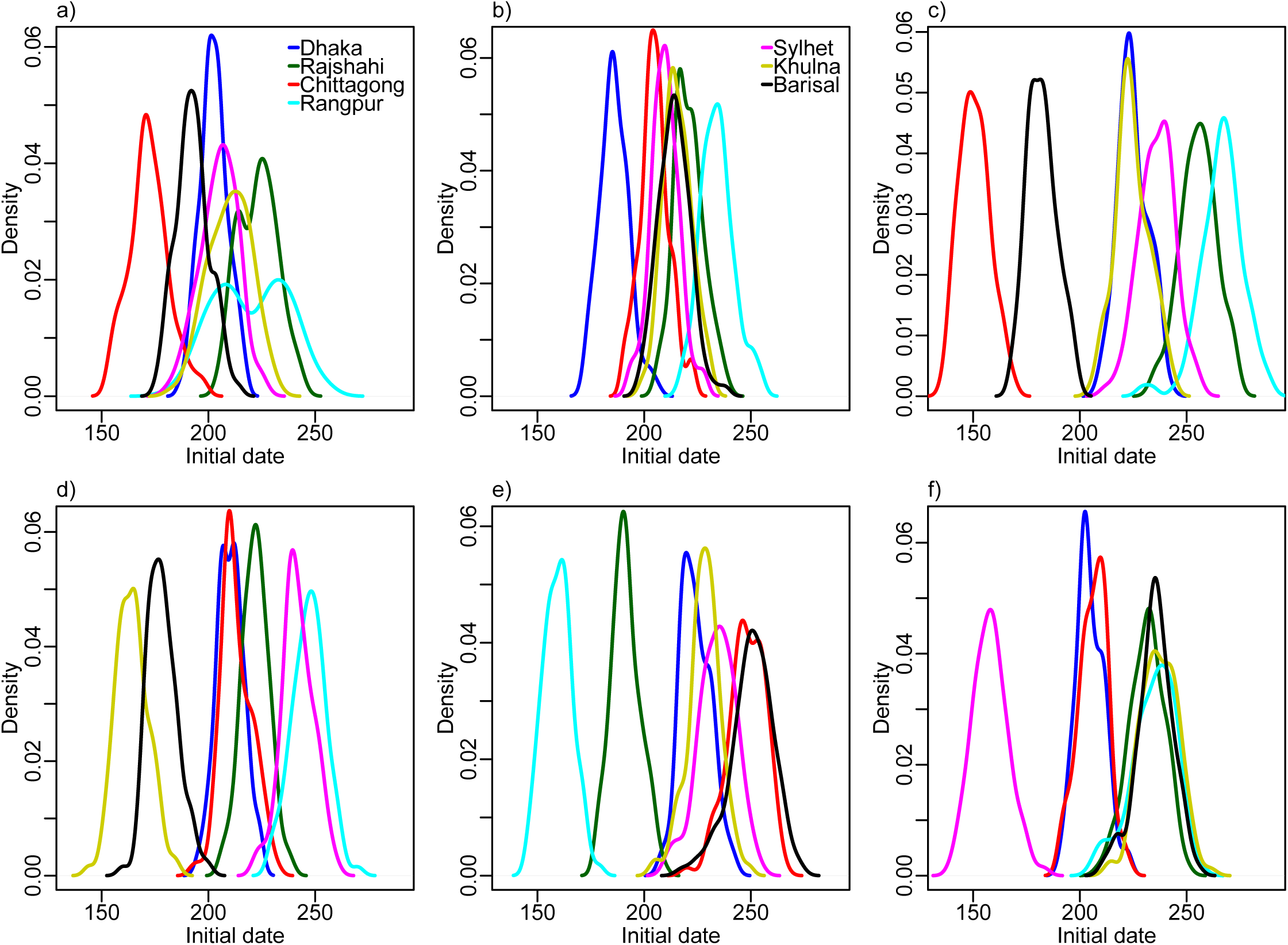
Radiation model. Initial dates distribution for each division for the radiation model, seeding in a) 2017 simulation, b) Dhaka, c) Chittagong, d) Khulna, e) Rangpur, and f) Sylhet.

**Figure 10:**
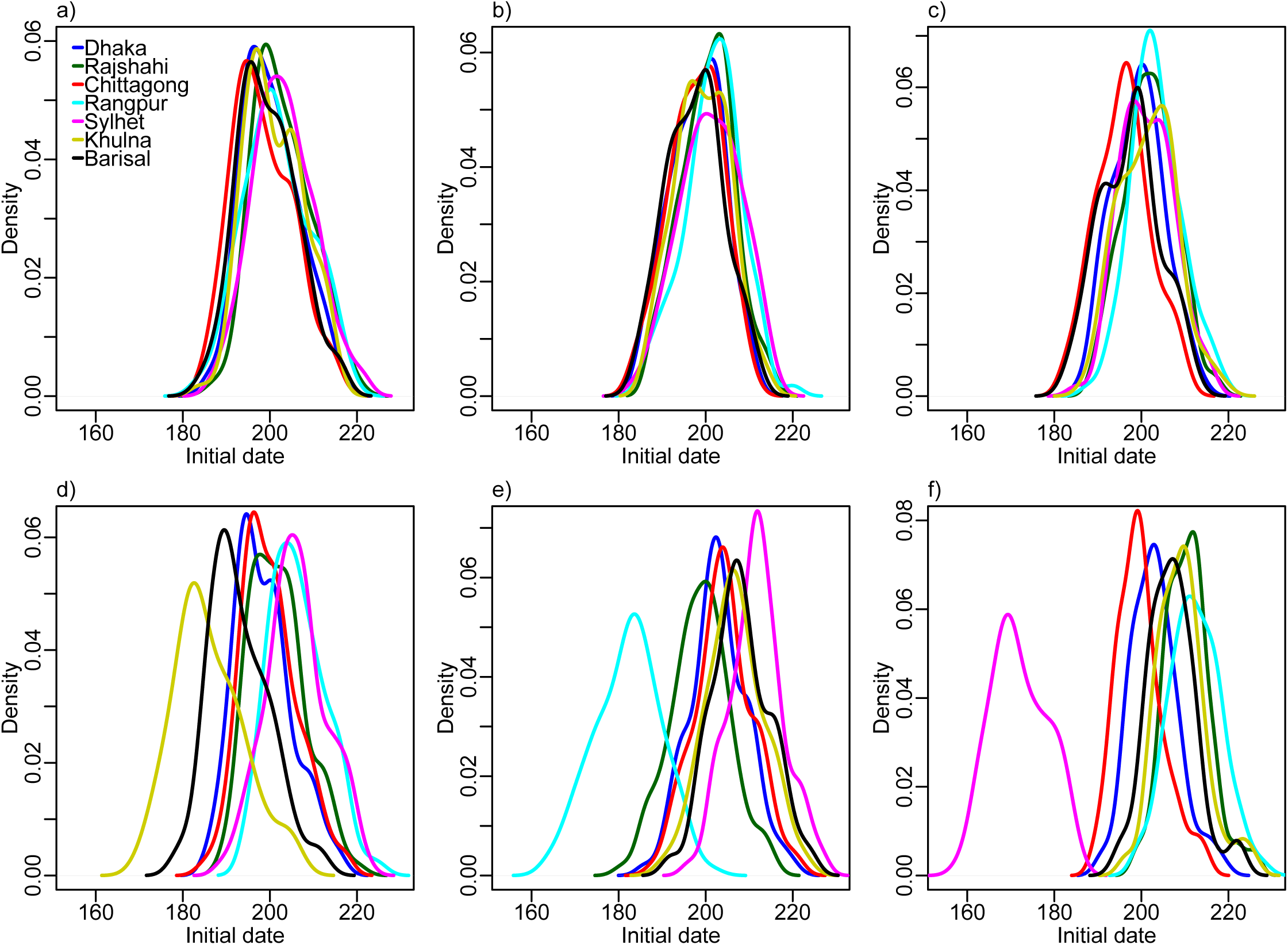
Time-averaged mobility. Initial dates distribution for each division for the time-averaged mobility, seeding in a) 2017 simulation, b) Dhaka, c) Chittagong, d) Khulna, e) Rangpur, and f) Sylhet.

**Figure 11:**
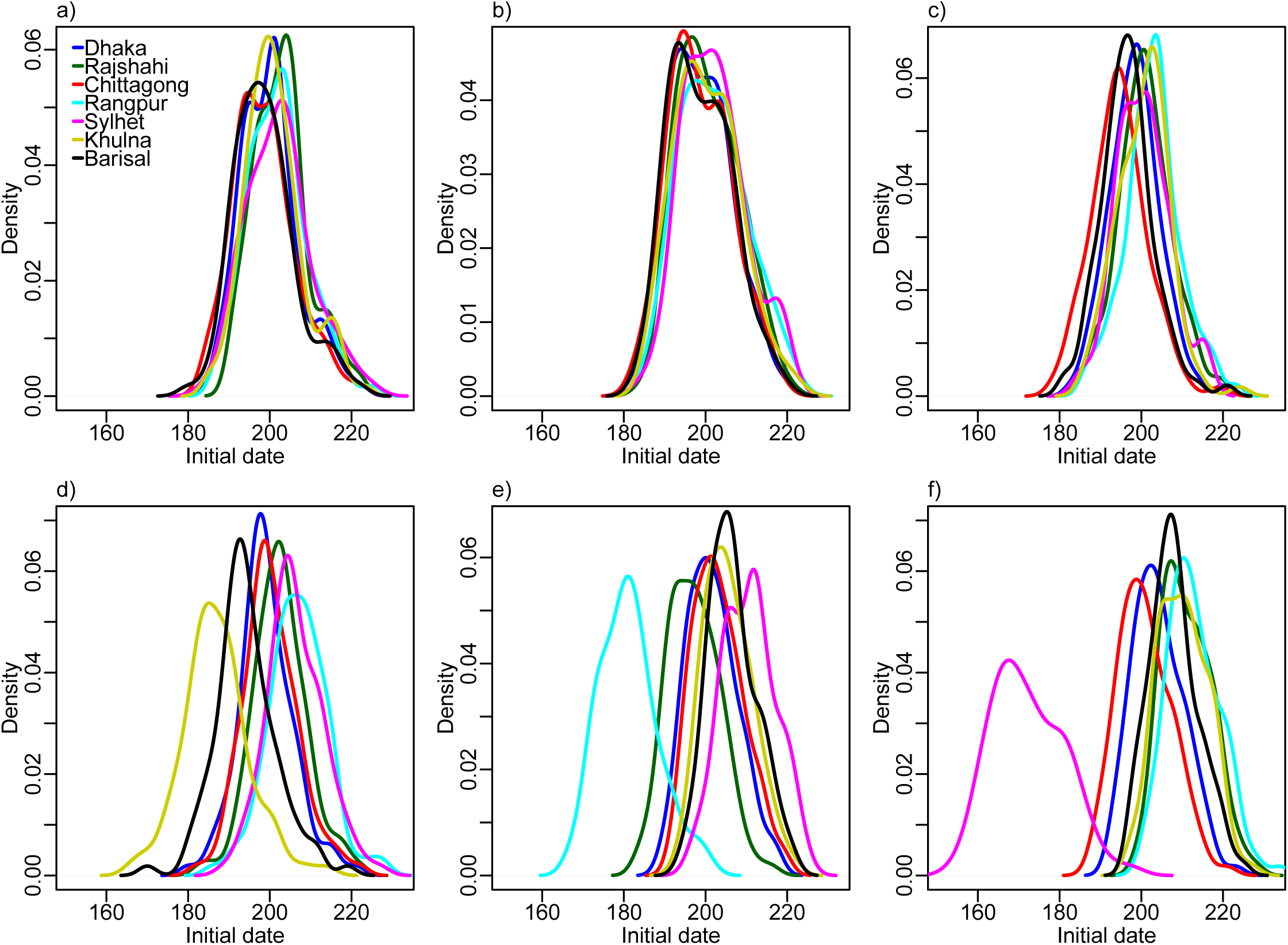
Daily mobility seeding on 1st of April 2017. Initial dates distribution for each division for the daily mobility when seeding on the 1st of April 2017, seeding in a) 2017 simulation, b) Dhaka, c) Chittagong, d) Khulna, e) Rangpur, and f) Sylhet.

**Figure 12:**
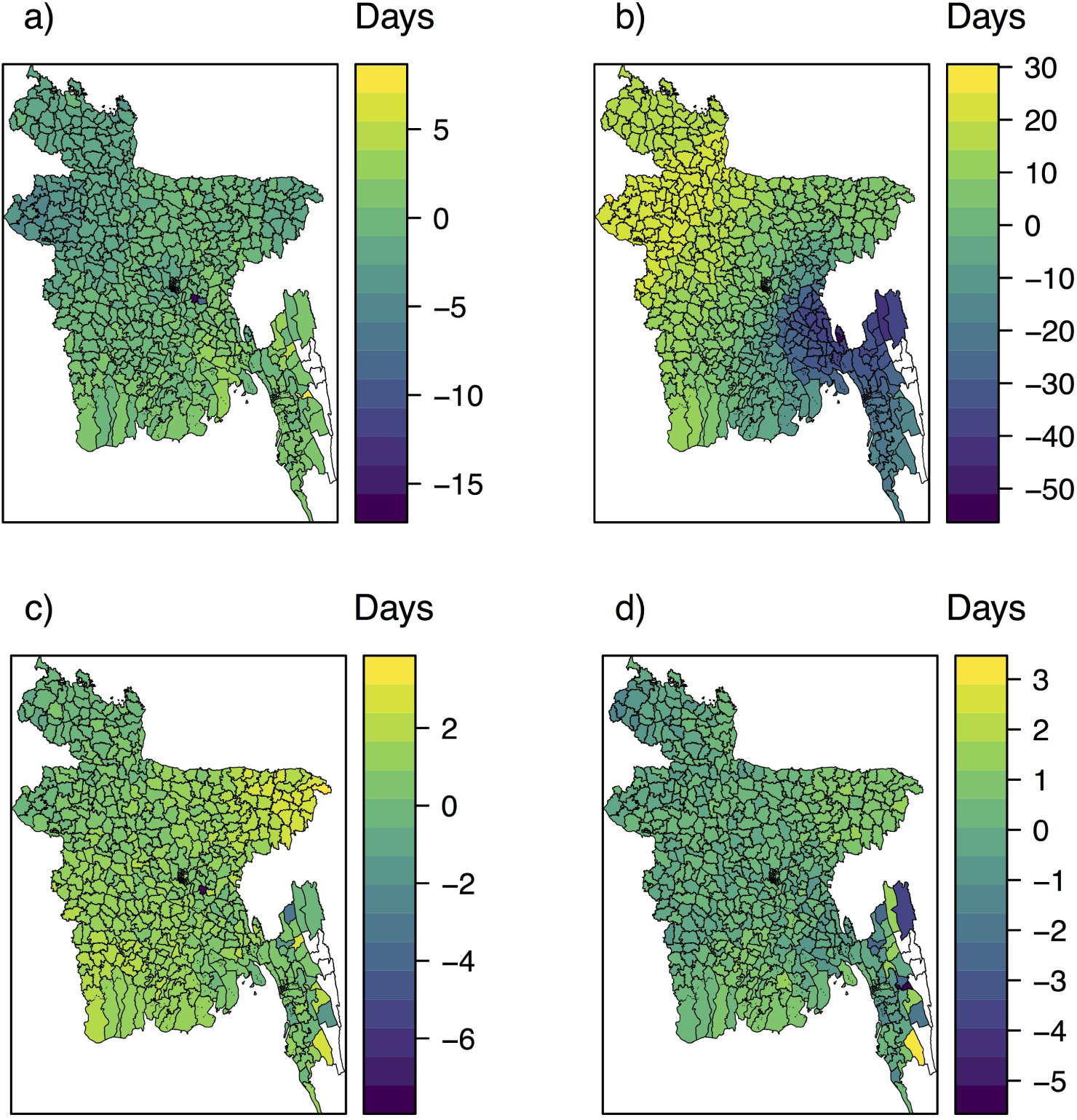
Initial dates difference between the mobility approximations and the daily mobility when seeding according to the 2017 season for a) the gravity model, b) the radiation model, c) the time-averaged mobility, and d) the time-averaged mobility when seeding on the 1st of April 2017.

The difference in initial dates between the time-averaged mobility model and the daily mobility model for each upazila is given in Figure 12c. Most of these values were positive, with a mean of 0.90 (std 0.82), indicating slightly delayed spread with the time-averaged mobility. The epidemic in the upazilas of the North-East was slightly delayed under the time-averaged mobility, while the upazilas in the North-West were hit slightly too early. The differences were much smaller than for the gravity and radiation models. The correlation between the mean initial dates under the time-averaged mobility and the daily mobility model was 0.96. The correlations between the initial dates under the mobility approximations and the daily mobility for the other seeding scenarios are provided in electronic supplementary material Section S9. The correlations with the time-averaged mobility were high, and higher than with the gravity and radiation models.

The difference in initial dates between the time-averaged mobility setting and the daily mobility setting when seeding on the 1st of April 2017 for each upazila is given in Figure 12d. The differences were small, with a mean of 0.12 (std 0.62). The correlation between the mean initial dates was high, 0.98.

In the electronic supplementary material Sections S8, S10, and S11, we plot the early spatial spread for the different seeding scenarios, by the prevalence at early time points. The spread was faster with the gravity model than with the daily mobility, and slower with the radiation model. For the radiation model, the early spread is mainly to proximate locations. The early spread using time-averaged mobility was similar to the early spread using daily mobility.

We assessed the relative performance of the radiation model and the gravity model on replicating the mobile phone mobility in electronic supplementary material Section S14. The radiation model had a higher correlation with the mobile phone data, but greatly overestimated the travel on some links. The radiation model performed overall best, but the gravity model outperformed the radiation model for short distances and travel between small and large population sizes. The density of the radiation mobility network was much lower than for the time-averaged mobility network, while the density of the gravity mobility network was much higher. This might explain why the gravity model overestimates the spatial synchrony and early spread, while the radiation model underestimates it.

### 4.3 Influenza in Bangladesh

The order of initial dates for the 2017 simulation was Chittagong, Barisal, Dhaka, Khulna, Sylhet, Rangpur, and Rajshahi. Similarly, in the case data, Dhaka division was reached first, followed by Chittagong and then Barisal (Figure 13).

**Figure 13:**
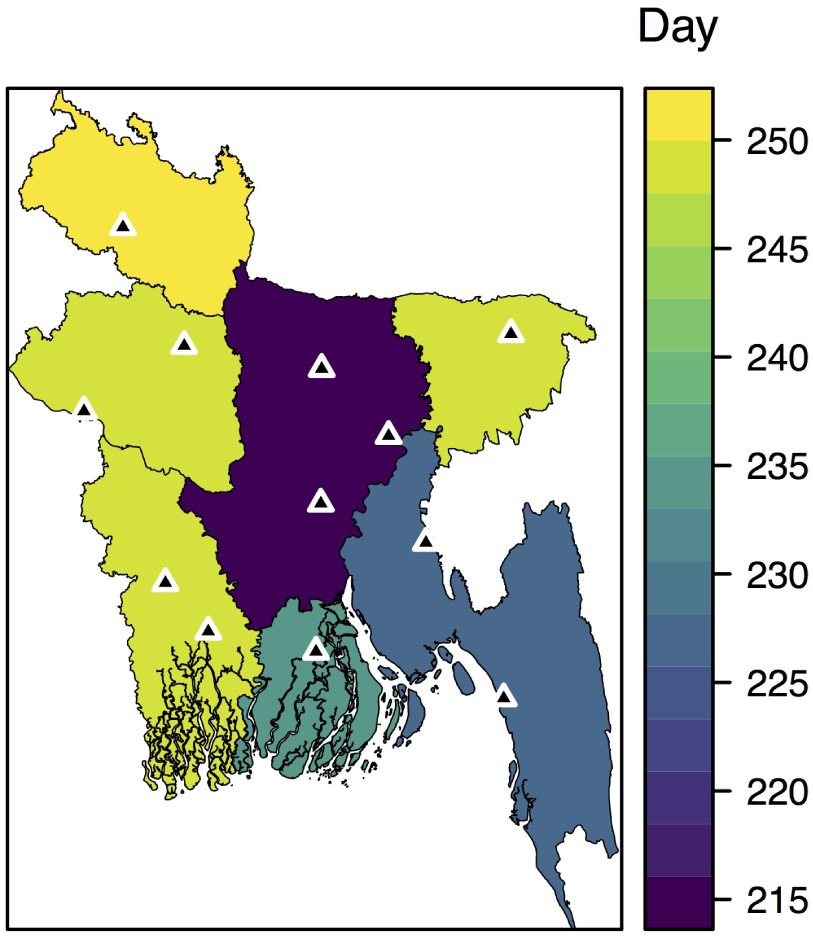
Arrival times on division level for the 2017 case data, defined as the day when 10% of the total number of cases have occurred. The sentinel hospitals are marked by black triangles.

The distribution of initial dates for the different divisions are given in Figures 7a-f, when seeding in early locations for 2017, Dhaka, Chittagong, Khulna, Rangpur, and Sylhet, respectively. The relative order of arrival is provided in Table 3. Regardless of seeding location, Chittagong and Dhaka seemed to be reached early. In general, Rajshahi, Rangpur, and Sylhet were hit later, unless we seeded in these divisions. The spatial spread was more coherent when seeding in Dhaka and Chittagong, than when seeding in Khulna, Rangpur, or Sylhet. Seeding in Dhaka, the delay was four days between the epidemic sparked in the latest and earliest hit division. The corresponding delay was approximately one week when seeding in Chittagong, three weeks when seeding in Khulna, four weeks when seeding in Rangpur, and six weeks when seeding in Sylhet.

The early spatial diffusion is provided in electronic supplementary material Section S8. The early spread seemed to be both radially from the seeding location, but also to larger cities, in particular the capital.

### 4.4 Other influenza seasons

The estimated transmissibility for the 2015 influenza season was lower than for the other seasons (Table 1). This coincides with the fact that this season had the lowest proportion of A(H3) cases. Seasons dominated by A(H1N1) and B have been found to be milder than seasons dominated by A(H3) (18). The transmissibilities for 2014 and 2017 were higher than for the other seasons. Note that the posterior credible intervals for *r* were overlapping, indicating that the data do not contain enough information on *r*. There was less overlap for *β*, and the posterior credible intervals were narrow.

The model had a good qualitative fit for the 2014-2016 seasons. The simulations and posterior distributions are provided in electronic supplementary material Section S15.

## 5 Discussion

### 5.1 Evaluation of mobility approximations

Using time-averaged mobile phone data resulted in a good approximation to the spatio-temporal disease dynamics in Bangladesh, projected by models informed by daily mobility data. Thus, this type of data appears to be a viable alternative to model human movements in the context of directly transmissible respiratory infections. Our finding is in accordance with studies of individual mobile phone mobility trajectories, showing that human mobility is highly predictable and regular in both time and space (19; 20; 21).

Both model-derived approximations to human movement resulted in significant loss of accuracy to predict the epidemic spread. The unconstrained gravity model, in particular, produced epidemics that rapidly leaped across the country, resulting in almost synchronous epidemic signals. The opposite tendency occurred with the radiation model. The dissemination was slower, evolving in forest-fire-like patterns. Consequently, the spatial synchrony and peak prevalence were underestimated. Our analyses suggest that the simplified gravity and radiation models are thus not detailed or accurate enough to capture all the relevant aspects of the information-rich mobile phone data. Similarly, (4) and (5) found that the gravity model was unsatisfactory compared to mobile phone mobility to model a cholera outbreak in Senegal and dengue transmission in Pakistan, respectively. (22) compared the use of gravity and radiation models to mobile phone mobility data in Kenya, and found that neither performed well, in particular for rural areas. Similar to our result, they found that the gravity model overestimated spatial travel.

The mobile phone data are aggregated and potentially biased. However, we use the daily mobility as the “gold standard”, as they represent our only observation of the true, underlying human mobility. We note that even though the daily mobility data are not perfect, there is little reason to expect that the time-averaged mobility would perform worse compared to daily mobility for higher quality mobility data.

### 5.2 Generalisability and alternatives for time-averaged mobility

We chose to use the time-averaged mobility for the days without daily information, and for the influenza seasons 2014-2016. This approach relies on two implicit assumptions. One is that the time-averaged mobility sufficiently describes the daily mobility, which we have justified. The other is that the mobility patterns do not change much from season to season. The model was able to capture the data for the various influenza seasons well. This further motivates and supports a more general use of time-averaged mobility. The time-averaged mobility tends to smooth the mobility network, because human flow between locations that are only realised particular days, in this approximation will be effective every day, but with a lower travel volume. An alternative to the use of time-averaged mobility would be to add noise to the time-averaged mobility matrix. Another alternative is to employ separate matrices for weekends and weekdays, or even one matrix per weekday. In (4), mobile phone data from 2013 were used to assess the effect of a mass gathering on a cholera outbreak in 2005 in Senegal. They found that none of their more detailed mobility approximations performed better than the time-averaged mobility matrix. This further justifies our parsimonious choice of using time-averaged mobility, instead of more detailed models including higher order effects like for instance weekends.

### 5.3 Privacy concerns and implications

Mobile phone data are sensitive and subject to privacy challenges (8). Human mobility is unique and thus also identifiable, and even coarse data sets are not fully anonymous (8). Our data are therefore aggregated in both time and space, and do not allow tracking trips that last for more than one day. We have found that for our setting, averaging mobility over multiple days is a good approximation to daily mobility. This is advantageous, as this matrix contains less temporal information on individual movement than the daily mobility matrices.

Aggregation leads to fewer privacy concerns. However, we have not formally tested the effect sizes, as it would require individual level data which we have not had access to. Future studies should compare uniqueness and re-identification of individuals for the daily mobility and the time-averaged mobility.

### 5.4 Implications for Bangladesh

Our model fits the case data well, considering the scarcity of the data. The ability to accurately predict epidemic spread in developing countries like Bangladesh has many useful implications for public health. In particular, insights into those spatio-temporal trends can improve pandemic preparedness planning. One quantitative important finding is the coherency in the predicted epidemic spread. When seeding in Chittagong or Dhaka, it took approximately a week from the epidemic sparked in the earliest division until it had sparked in all divisions, and hence there is little time to implement targeted interventions if the epidemic starts in Chittagong or Dhaka. However, when the epidemic started in one of the other divisions, the spread was less coherent, and the corresponding time ranged from three to six weeks, with more time to implement targeted interventions. The results when we seed in one, single upazila are likely to be more relevant for epidemics that start locally, like for instance avian influenza, than for an imported epidemic, which would likely have multiple seeding events.

Our results are in agreement with (23), who found that the 2007 seasonal influenza epidemic likely started in Chittagong and Dhaka. We find that Chittagong and Dhaka experienced early initial dates, regardless of seeding location. Though the scarce case data limit our ability to assess how well our model captures the spatial spread, the relative arrival times of our model in the different divisions are in good agreement with the data, with Dhaka, Chittagong, and Barisal being hit before the other divisions.

Our estimates of *R*_*e*_ range from 1.15 to 1.22. In a systematic review, (24) found a median value of *R*_*e*_ for seasonal influenza of 1.28 with an IQR of 1.19 to 1.37, hence our estimates lie in the lower range. Variations in the reproductive number between different populations are expected, due to for instance varying contact rates, age structure, humidity, and other climatic factors (24). Local estimates of the reproductive number are important, as small variations can be crucial for determining the success of a control measure in containing the epidemic. Moreover, local estimates of the reproductive number for seasonal influenza in tropical countries are needed to determine if and how transmissibility depends on geographical and social factors (24).

### 5.5 Limitations

#### 5.5.1 Limited case data and forecasting

The limited spatial information in the case data prevents us from formally assessing which mobility model best captures the actual disease spread. Though the sentinel surveillance platform is comprehensive and very valuable for public health purposes, etc, spatial mathematical models of transmission perform best with more spatial granularity and a larger volume.

The case data are not rich enough to assess forecasting accuracy. This would require estimating the parameters based on only a few weeks of data, and given our data volume, the credible intervals would be very wide. In addition, the summary statistics we have used in our ABC-SMC procedure cannot be used early in the epidemic. An alternative summary statistic in such settings is the early growth rate. The model can, however, be used to predict relative arrival times in different regions, since they are not very sensitive to the exact epidemiological parameters (25).

#### 5.5.2 Observation process

We assume that the number of observed cases follows a binomial distribution with a constant reporting probability. It might however be that a beta-binomial observation model is more suitable, due to high variance. We have chosen to use the simpler binomial model due to the scarce case data, as the beta-binomial distribution requires estimation of an additional parameter. In addition, the reporting rate might depend on the process, i.e. temporal dependence through the infectious disease activity. The detection probability for the surveillance system has been found to decrease with distance to the hospitals (26). In addition, healthcare seeking depends on socio-economic status (26), which can also be space dependent. However, the 12 hospitals are spread out throughout the country. We have chosen to use a constant reporting probability, due to scarce data. Moreover, these choices are computationally more efficient, as fewer parameters have to be estimated. Computational cost is a limitation of our fitting procedure–fitting the model for one influenza season took approximately one week running in parallel on 112 cores.

#### 5.5.3 Mobility assumptions

As we do not have individual identifiers in the data, we choose to let individuals travel in the morning according to the mobile phone mobility data, and send them back to the home location after 24 hours. In this way, we do not allow trips which last for more than 24 hours. We choose to model travel like this, as it is reasonable to assume that individuals preferentially return to their home location, since human mobility is characterised by preferential return and reproducible patterns (19; 21). Moreover, humans have a tendency to return home on a daily basis (21). Alternatively, one could have let individuals move permanently to their destination location. However, this would overestimate the synchrony between the locations, and could result in very varying population sizes. In addition, since we do not have individual identifiers, the travellers are randomly drawn from the home population. In reality, there is likely some regularity in who travel on the same link on different days, as human mobility is characterised by preferential return to a few locations (19; 21). The fact that we cannot capture regular movements is likely to overestimate the spread (27).

#### 5.5.4. Bias in mobile phone data

A potential source of bias is the mobile phone data, and whether the population captured in the data is representative of the Bangladeshi population. As of June 2017, 54% of the Bangladeshi population subscribed to mobile services (28). Grameenphone is the largest mobile operator in Bangladesh, but there may be market share bias. Young children typically do not own a mobile phone. Women in Bangladesh are 32% less likely to own a mobile phone than men (28). For Kenya, mobile phone mobility estimates were found to be robust to ownership bias (29). To the extent that these results can be translated to Bangladesh, the mobile phone mobility estimates are a good proxy for human mobility, despite the ownership bias. A good agreement between commuting networks based on mobile phone data and census data was also found in (30), for Spain, Portugal, and France.

The mobile phone data are however likely to be subject to overestimation bias, since the population who travel most frequently are those with fewest economic constraints, and thus more likely to own a mobile phone (29). Overestimation of mobility might overestimate the spatial synchrony of the epidemic. It is, however, not likely to affect the relative arrival times in the locations, given that the mobile phone mobility is otherwise unbiased (i.e. the overestimation is constant in space). For example, (30) found that mobile phone data overestimated commuting, but that the overestimation did not greatly affect the order of arrival times in different spatial regions. Travel surveys, on the other hand, have been found to underestimate human mobility (31). We claim that underestimating human mobility is a more severe problem, resulting in underestimation of spatial synchrony, and thus too optimistic estimates of how much time one has to implement targeted interventions. Moreover, we only have mobile phone data for six months, and we thus do not capture seasonal variations.

Another limitation is that we do not have information on spatial dependence of mobile phone subscriptions. We have therefore assumed uniform coverage when we estimate the population sizes and the mobility matrices from the mobile phone data. Alternatively, the population sizes could have been estimated by use of satellite imagery (32).

#### 5.5.5 Aggregation scale

The temporal aggregation scale is 24 hours, hence regular commuting and short-range travel are not well captured. The mobile phone data are inherently noisy, since the user’s location is only captured when a service takes place (7). Consequently, a location change can fail to be captured if there is no phone activity, and the estimated location for a given day might not correspond to the location where they spent the most time. The spatial scale is a trade-off between capturing relevant signal and details, computational intensity, and aggregating out noise. With a high resolution, the accuracy of the mobility estimates depends on cell tower density, individual calling behaviour, and frequency, which are sources of noise and bias (33). Aggregated mobility is less biased by these factors (33). Moreover, a coarser spatial resolution means less privacy concerns. However, a finer spatial scale makes the homogeneous mixing assumption more reasonable, and captures more heterogeneity and details. The computational cost scales with the number of compartments and links, and for each additional location, there are five additional compartments and many links. For convenience, we use administrative units. The influenza case data are provided on the scale of administrative units. For computational efficiency, we have chosen to use the upazila scale, as a finer resolution would be very computationally expensive, and district level would be too coarse.

#### 5.5.6 Bias in influenza data

Hospital data with confirmed cases are less noisy than for instance general practitioner data, which are subject to varying healthcare seeking behaviour and noise due to other respiratory infections. For fatal respiratory infections in Bangladesh, (26) found that elderlies were under-represented in surveillance cases, while children under 5 years were overrepresented, compared to the cases in the population. This is a potential source of bias in the temporal pattern. Children have been hypothesised to drive local transmission, while adults spread the disease to new locations, resulting in shifted epidemic timings for different age groups (34). This effect can also interact with the specific seasonal influenza strain, as some seasons are associated with cases among the elderly, while others are associated with more and earlier cases among children (34).

### 5.6 Contributions and future perspectives

Our study is a contribution to understanding the detail level of mobile phone mobility data needed for modelling infectious diseases, with important privacy-conserving consequences. We further contribute to understanding how the gravity and radiation models perform in a developing country. We provide estimates of the relative arrival time in the different divisions for various seeding scenarios, with important preparedness planning implications. We obtain the first local estimates of the reproductive number for seasonal influenza in Bangladesh. Our example shows the use of ABC in a setting with scarce influenza case data, which can contribute to motivate the use of rigorous statistical methods in infectious disease modelling.

The fact that the time-averaged mobility was a good approximation motivates a more general use–this is an encouraging result for outbreak control and future models, as daily mobile phone mobility is unlikely to be available in real-time during an outbreak. This is particularly valuable for low-income settings, where census data are scarce. Future studies should assess the generalisability of time-averaged mobility to future outbreaks, by investigating whether the essential mobility patterns for infectious disease transmission stay relatively constant over time.

Our study is a contribution to using novel data sources in infectious disease modelling, utilising mobile phone data as a proxy for human mobility. This is especially useful for countries with little or low-quality census data. In recent years, novel data streams have been increasingly popularised in disease surveillance and prediction (35). These data sources are often noisier than traditional surveillance data sources, but can improve timeliness, spatial and temporal resolution (35). In the future, it would be interesting to combine the good signal, low-volume hospital sentinel data with large-volume novel data sources, to inform the model parameters and assess the spatial spread, for instance by exploiting search queries, social media like Twitter, or

Wikipedia access logs. Incorporating multiple data sources in ABC is straightforward.

## Data Availability

All requests for the mobile phone dataset should be directed to Kenth Engø-Monsen at Telenor Research. Requests for the influenza data should be directed to Mohammad Abdul Aleem at icddr,b.

## 6 Acknowledgement

icddr,b is grateful to the governments of Bangladesh, Canada, Sweden and the UK for providing core/unrestricted support. The manuscript has been reviewed by icddr,b for scientific content and consistency of data interpretation with previous icddr,b publications.

## 7 Funding

SE and AF acknowledge partial funding from the Norwegian Research Council centre BigInsight project 237718.

## Notes

### Competing Interest Statement

The authors have declared no competing interest.

### Funding Statement

icddr,b is grateful to the governments of Bangladesh, Canada, Sweden and the UK for providing core/unrestricted support. SE and AF acknowledge partial funding from the Norwegian Research Council centre BigInsight
project 237718.

